# Prespecified Internal Pilot and Feasibility Framework for a Pragmatic Randomized Controlled Trial Comparing Mechanical Diagnosis and Therapy Versus Generalized Exercise in Surgeons With Chronic Spinal Pain: A Protocol

**DOI:** 10.64898/2026.04.08.26350288

**Authors:** Claus Kjærgaard, Pascal Madeleine, Annett Dalbøge, Benjamin Steinhilber, Anne Vingaard Olesen, Tommy Kjærgaard Nielsen

## Abstract

**Background:** Trials in occupational populations, such as surgeons, face feasibility challenges due to high workload, restricted availability, and clinical heterogeneity, which may compromise recruitment, adherence, and retention.

**Objective:** To prespecify the feasibility framework and progression criteria for an internal pilot phase embedded within a pragmatic randomized controlled trial (RCT) comparing Mechanical Diagnosis and Therapy with generalized exercise in surgeons with chronic spinal pain.

**Design:** Protocol for a prespecified internal pilot phase embedded within a pragmatic, two-arm, parallel-group RCT.

**Methods:** The internal pilot will include the first four months of recruitment and aims to randomize at least 12 participants. Feasibility will be assessed across predefined domains, i.e., recruitment, eligibility, consent, intervention uptake, adherence, retention, data completeness, and treatment fidelity. Each domain is operationally defined and linked to prespecified progression criteria to ensure interpretability and decision-making utility. Criteria will be interpreted collectively to guide trial continuation. A minimal qualitative process evaluation will be embedded.

**Ethics and dissemination:** The host trial has received ethical approval (N-20240046) and is registered at ClinicalTrials.gov (NCT07293130). The findings from the internal pilot will be reported in a separate feasibility manuscript.

## INTRODUCTION

### Background

Trials in occupational populations, such as surgeons, face substantial feasibility challenges due to high workload, limited availability due to on-call shifts and rotating shift schedules, and clinical heterogeneity, which may compromise recruitment, adherence, and retention [4]. Feasibility studies are conducted to determine whether a future randomized controlled trial (RCT) can be performed, whether it should proceed, and how it should be optimized [1, 2]. Internal pilot studies represent a specific design in which feasibility is assessed within the early phase of the definitive trial, allowing for participant retention and improved efficiency [3]. Chronic spinal pain is highly prevalent among surgeons and is associated with sustained static postures and occupational demands [5]. Exercise-based interventions are recommended. However, these have shown modest average effects, likely reflecting heterogeneity in treatment response [6-8].

Mechanical Diagnosis and Therapy (MDT) is a classification-based approach that targets directional preference and may improve outcomes in selected subgroups [9-11]. However, their effectiveness must be evaluated within a feasible trial design. Therefore, this study prespecifies an internal pilot phase and feasibility framework embedded within a pragmatic RCT comparing MDT with generalized exercise.

## METHODS

### Study design and host trial context

This study describes a prespecified internal pilot phase embedded within a pragmatic, two-arm, parallel-group randomized controlled trial with 1:1 allocation.

The host trial includes surgeons aged 25–70 years with chronic spinal pain (>3 months) and a confirmed directional preference. Participants are randomized to MDT or generalized exercise. The primary outcome of the host trial is the average spinal pain intensity over the preceding week (Numeric Pain Rating Scale, 0–10) at 12 weeks.

Participants included during the internal pilot will be retained in the main trial.

### Internal pilot phase

The internal pilot will include the first four months of recruitment and aims to randomize at least 12 participants to support interpretable estimation of feasibility outcomes. If fewer than 12 participants are randomized within this period, feasibility outcomes will still be summarized descriptively and interpreted cautiously, with particular emphasis on recruitment barriers and implications for continuation of the host trial. The internal pilot duration is fixed to ensure timely feasibility assessment independent of recruitment performance.

The duration and minimum sample size are chosen pragmatically to allow an early but informative assessment of key feasibility parameters, particularly the recruitment rate, adherence, and retention. This timeframe is expected to capture initial recruitment dynamics and early intervention engagement while enabling timely adjustments to trial procedures if needed. Effectiveness analyses will not be conducted during the pilot phase.

### Feasibility framework

Feasibility will be assessed across domains recommended for pilot studies [1, 2, 12], with each domain operationally defined and directly linked to progression criteria to ensure interpretability:

- Recruitment rate: Number of randomized participants per month
- Eligibility proportion: Proportion of screened individuals meeting inclusion criteria
- Consent rate: Proportion of eligible participants randomized
- Intervention uptake: Proportion initiating ≥1 session within 3 weeks
- Adherence: Proportion completing ≥3 sessions within 12 weeks
- Retention: Proportion completing 12-week follow-up
- Data completeness: Proportion with primary outcome at 12 weeks
- Treatment fidelity: Proportion delivered according to protocol

### Data sources

Feasibility data will be obtained from screening logs, enrolment logs, physiotherapy logs, REDCap, and clinical record audits. Reasons for non-participation and dropout will be recorded where available to support interpretation of potential selection and attrition bias.

### Progression criteria

Prespecified progression criteria are defined for each domain (Table 1) and will not be modified after inclusion of the first participant.

**Table 1.** Prespecified progression criteria for the internal pilot phase.

| Domain | Metric | Green \| Amber \| Red |  |  |
| --- | --- | --- | --- | --- |
| Recruitment | Randomized/month | $\geq 3$ | 2.0–2.9 | <2 |
| Eligibility | Eligible/screened | $\geq 35\%$ | 20–34% | <20% |
| Consent | Randomized/eligible | $\geq 60\%$ | 40–59% | <40% |
| Intervention uptake | $\geq 1$ session within 3 weeks | $\geq 85\%$ | 70–84% | <70% |
| Adherence | $\geq 3$ sessions by 12 weeks | $\geq 75\%$ | 60–74% | <60% |
| Retention | 12-week follow-up | $\geq 85\%$ | 75–84% | <75% |
| Data completeness | NPRS at 12 weeks | $\geq 90\%$ | 80–89% | <80% |
| Fidelity | Protocol concordance | $\geq 80\%$ | 65–79% | <65% |

Consent: Provision of written informed consent followed by randomization among eligible participants; Adherence: Attendance at ≥3 supervised treatment sessions within 12 weeks, unless earlier discharge occurs due to goal achievement; NPRS: Numeric Pain Rating Scale

Progression criteria will be interpreted collectively and in relation to each other, rather than in isolation, acknowledging that feasibility domains are interdependent:

- Green: Continue as planned
- Amber: Continue with targeted adjustments
- Red: Initiate structured review and corrective actions

This approach reflects recommendations that progression criteria should guide, rather than dictate, decision-making [1-3]. Given the absence of universally accepted cut-offs, thresholds are selected pragmatically, informed by feasibility trial guidance and expected recruitment capacity, adherence, and retention in this occupational population.

### Decision-making process

Decisions regarding continuation or modification will be made by the investigator group, including the principal investigator and senior investigators, and documented in a decision log to ensure transparency.

Potential corrective actions include expansion of recruitment sites, adaptation of intervention delivery, and enhanced follow-up procedures.

### Process evaluation

A minimal qualitative process evaluation will be embedded to support the interpretation of feasibility outcomes. Semi-structured interviews will be conducted with a purposive sample of participants (anticipated n=3 to 6), depending on recruitment and data sufficiency. Sampling will aim to include participants with varying levels of adherence.

Interviews will be conducted via telephone or video consultation, audio-recorded where feasible, and summarized using structured field notes. Data will be analyzed using a pragmatic thematic approach to identify key barriers, facilitators, and suggestions for improvement. The findings will be used to contextualize feasibility outcomes and inform potential trial modifications.

### Documentation of deviations

All deviations from protocol procedures, feasibility definitions or thresholds, or planned corrective actions, will be recorded in a prospectively maintained decision log. This log will include date, issue, action taken, and rationale. Any modifications to the trial procedures will not involve changes to the prespecified feasibility definitions, outcome measures, or progression thresholds.

### Statistical analysis

Feasibility outcomes will be summarized descriptively using proportions with 95% confidence intervals. No hypothesis testing or between-group effectiveness analyses will be conducted. The findings will be interpreted against the prespecified progression criteria. Given the small sample size, estimates of feasibility outcomes are expected to be imprecise, and confidence intervals will be interpreted cautiously.

### Ethics and dissemination

The host trial has been approved by the North Denmark Region Committee on Health Research Ethics (N-20240046) and registered at ClinicalTrials.gov (NCT07293130). All participants provide informed consent.

Feasibility findings will be reported descriptively and interpreted against prespecified criteria. The results will be disseminated through peer-reviewed publications.

## DISCUSSION

This prespecified internal pilot framework enhances transparency and reduces the risk of post hoc decision-making and selective interpretation. Internal pilot designs are increasingly recommended to improve trial efficiency, particularly in settings where recruitment, adherence, and retention are anticipated challenges [3]. Embedding feasibility assessments within host trials reduces duplication of effort and contributes to minimizing research waste.

Recruitment through professional networks and voluntary participation may introduce selection bias, potentially resulting in the overrepresentation of highly motivated participants and overestimation of adherence and retention. In addition, restricting inclusion to participants with a directional preference limits generalizability and may affect recruitment feasibility. These factors should be considered when interpreting feasibility outcomes.

Feasibility estimates derived from early recruitment phases may be influenced by startup effects and early adopters, potentially overestimating feasibility. The small sample size is expected to result in imprecise estimates and should be interpreted cautiously.

A key strength of this approach is the integration of quantitative feasibility metrics with a qualitative process evaluation, enabling both measurement and contextual understanding of feasibility challenges.

The prespecified feasibility domains are directly linked to the viability of the definitive trial. Recruitment and consent rates determine the ability to achieve the planned sample size, whereas adherence, retention, and data completeness influence the validity and interpretability of trial outcomes. Treatment fidelity is essential to ensure that the intended intervention contrast is maintained. Collectively, these domains provide a structured basis for determining whether the trial can proceed as planned or requires modification.

Findings from the internal pilot will be used to inform targeted modifications to trial procedures where necessary, while preserving the integrity of the prespecified design.

## Conclusion

The internal pilot will provide prespecified, transparent, and structured evidence on feasibility across key domains and inform decisions regarding continuation and optimization of the definitive RCT.

## Data Availability

No datasets were generated or analyzed for this protocol. Data arising from the study will be made available upon reasonable request to the corresponding author, subject to applicable data protection regulations.

